# Self-assessment of problem-solving skills and Clinical Reasoning among sixth-year medical students University of Baghdad / College of medicine

**DOI:** 10.1101/2023.01.16.23284617

**Authors:** Ahmed M. Hasan, Ibrahim N. Abbas, Ahmed A. Abdulrazak, Ali Almothaffar

## Abstract

**Objectives:** To assess the level of problem solving skills of Sixth grade medical students of College of Medicine/ University of Baghdad using self-assessment tool.

**Materials and Methods:** A cross-sectional study to assess Problem Solving Disposition in Sixth year Medical Students in College of Medicine/ University of Baghdad. A printed survey of the questionnaire was distributed to sixth year medical students in august 2022. A sample of 151 students participated in the study by filling of the printed questionnaire which was validated by a previous study in a medical school in Mexico.

**Results:** The number of sixth Grade medical students in University of Baghdad/ College of medicine that participated in the study was 151 of whom 70 (46.35 %) were males and 81 (53.65 %) were females.

The mean score of the “knowledge of discipline” was 2.973± 0.999, as for the “pattern recognition” component the mean was 2.198 ±0.559, and the mean for the “application of general strategies” component was 2.158± 0.492.

The independent sample T-test results for the “Knowledge of discipline” category showed a mean score for females of 3.137 and a mean score for males of 2.778 with a p value >0.05.

For correlations between the categories one of the items of the “Pattern Recognition” category had a positive correlation with a statistical significance with 2 items from “Application of general strategies for problem solving” category.

**Conclusion:** Sixth year medical students in University of Baghdad/ college of medicine are better self-perceived in the pattern recognition and application of general strategies for problem-solving categories than the knowledge of discipline, males assess themselves in remembering concepts as better than their female counterparts.

## Introduction

A great deal of knowledge and skill is required to practice as a doctor. Physicians in the 21st century need to have a comprehensive knowledge of basic and clinical sciences, have good communication skills, and be able to perform procedures, work effectively in a team and demonstrate professional and ethical behavior. But how doctors think, reason and make decisions is arguably their most critical skill. [1]

Excellence in medicine is not just about good knowledge, skills and behaviors. While medical schools and postgraduate training programs teach and assess the knowledge and skills required to practice as a doctor, few offer comprehensive training in clinical reasoning or decision-making. This is important because studies suggest that diagnostic error is common and results in significant harm to patients. Diagnostic error typically has multiple causes, but two-thirds of the root causes involve human cognitive error – most commonly, when the available data are not synthesized correctly.

While some of this is due to inadequate knowledge, a significant amount is due to inadequate reasoning. [2]

Medical students master an enormous body of knowledge, but lack systematic problem solving ability and effective clinical decision making. [3]

Knowledge is not a collection of facts, but rather an ongoing process of examining information, evaluating that information, adding to it and reorganizing it, in order to solve a problem and make a diagnosis. It is like capital. Acquiring it isn’t sufficient but knowing how to invest and employ it in different circumstances is crucial [4].

Doctors are expected to make effective decisions in a well-defined manner in their medical career which necessitate the developing of critical thinking and problem solving skills strategies.

Critical thinking can be defined as the ability and willingness to assess claims and make objective judgments based on well-supported reasons. It is the ability to look for flaws in arguments and resist claims that have no supporting evidence. It also fosters the ability to be creative and constructive in generating possible explanations for findings, thinking of implications, and applying new knowledge to a broad range of social and personal problem [5]

Critical thinking involves asking questions, defining a problem, examining evidence, analyzing assumptions and biases, avoiding emotional reasoning, avoiding oversimplification, considering other interpretations, and tolerating ambiguity. Dealing with ambiguity is also recognized as an essential aspect of critical thinking. Ambiguity and doubt are necessary and even a productive part of the critical thinking process. [6]

Health care institutions are liable and prone to medial errors mainly diagnostic and management errors. Approximately one third of patient problems are mismanaged because of diagnostic errors. Part of the solution lies in improving the diagnostic skills and critical thinking abilities of physicians as they progress through medical school and residency Prevention of diagnostic errors is more complex than building safety checks into health care systems; it requires an understanding of critical thinking and clinical reasoning [7]

Medical problem-solving skills are essential to learning how to develop an effective differential diagnosis in an efficient manner, as well as how to engage in the reflective practice of medicine. Competency of a doctor is closely related to his critical thinking which he uses to solve daily problems and overcome it. A good doctor must be a good critical thinker and a good problem solver Competence can be defined as the habitual and judicious use of communication, knowledge, technical skills, clinical reasoning, emotion, values, and reflection in daily practice for the benefit of the individual and the community being served. [8]

In this research we want to assess the ability to solve problems in final year medical students since many of these skills have been confronted, tested and experienced throughout the medical school [9]. The Individual Generic Skills Test. A test that was developed by faculty members of a medical school in Mexico to assess the disposition of higher education students on diverse instrumental competencies.

The complete instrument measures self-perception on information literacy, problem solving, time management, self-direction, decision making, and critical thinking. [10]

Ability to solve problems has many components and skills that are incorporated to get a desired outcome. For instance, Norman and Schmidt proposed a three-category model for problem solving ability: (1) acquisition of factual knowledge, (2) mastery of general principles that can be transferred to solve new similar problems, and (3) pattern recognition. [10]

The purpose of this paper is analysis of students’ self-assessment of these skills so that they can improve their critical thinking abilities, know their strengths and improve it and find and overcome their weaknesses so they can be safe doctors for the community.

## Subjects and Methods

The study design is quantitative, descriptive, and non-experimental. The methodology used throughout the paper mimics and improves on the one used by the Mexican team in their research paper titled “Self-Assessment of Problem Solving Disposition in Medical Students”. [10]. It consists of two main phases: (1) applying the questionnaire which was already validated by the Mexican research team, and (2) analyzing the result with additional tools to extract more useful information.

### (1) Applying the questionnaire

The questionnaire [11] had three main categories which are: (a) Application of general strategies for problem-solving, (b) pattern recognition, and (c) knowledge of the discipline. Each had 4,2,1 questions in this order making the total questionnaire containing 7 items in total regarding the scientific subject of our paper plus a slot to identify their gender. The answering method used was a five choices Likert scale considers values closer to 1 as more favorable responses being closer to strongly agree and vice versa. This then was distributed to a group of (151) sixth-grade medical students at Baghdad University, College of medicine in August 2022. The students were asked to answer 7 questions in total each question relating to component in the problem solving disposition model, 1 question in the “knowledge of discipline” component, 2 questions in the “pattern recognition” component and 4 question in the “application of general strategies” component, and answers were obtained on a Likert scale ranging from 1 to 5, strongly agree being 1 and strongly disagree being 5 and agree and disagree assuming the values 2 and 4 respectively and neutral (undecided) being 3. See (Table1).

**Table 1:**
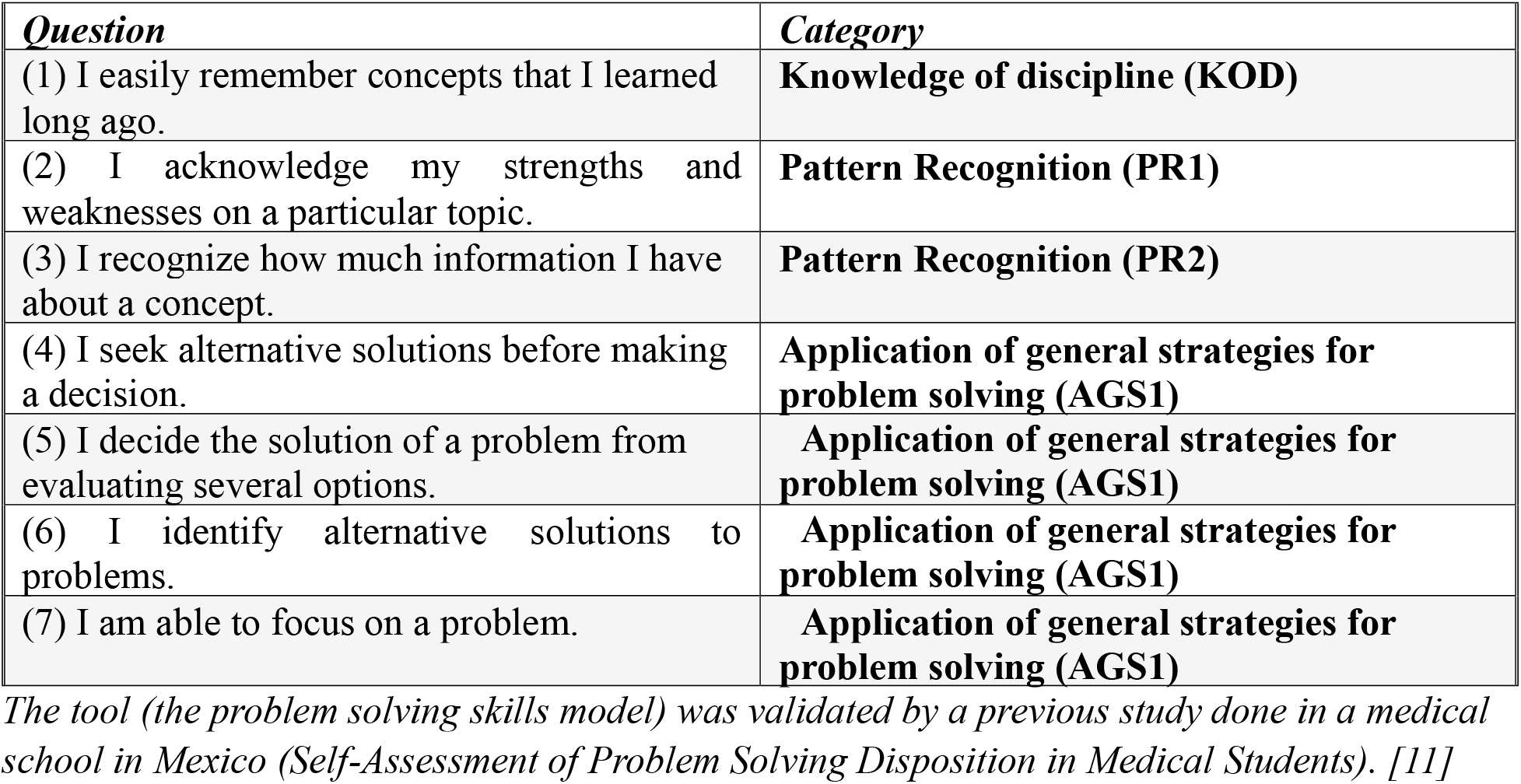
Items of the problem-solving skills model.

| ***Question*** | ***Category*** |
| --- | --- |
| (1) I easily remember concepts that I learned long ago. | **Knowledge of discipline (KOD)** |
| (2) I acknowledge my strengths and weaknesses on a particular topic. | **Pattern Recognition (PR1)** |
| (3) I recognize how much information I have about a concept. | **Pattern Recognition (PR2)** |
| (4) I seek alternative solutions before making a decision. | **Application of general strategies for problem solving (AGS1)** |
| (5) I decide the solution of a problem from evaluating several options. | **Application of general strategies for problem solving (AGS1)** |
| (6) I identify alternative solutions to problems. | **Application of general strategies for problem solving (AGS1)** |
| (7) I am able to focus on a problem. | **Application of general strategies for problem solving (AGS1)** |

*The tool (the problem solving skills model) was validated by a previous study done in a medical school in Mexico (Self-Assessment of Problem Solving Disposition in Medical Students). [11]*

### (2) Analyzing the results

after collecting the answers from our sampled population. The Statistical Package for the Social Sciences (SPSS, version 20) was used for the data analysis. We extracted the basic descriptive statistics for the total sample and each gender respectively (mean and standard deviation) plus all the frequencies for each item and category in the questionnaire.(See Table 6,7) An exploratory factor analysis was done to identify how many categories and to which each item belongs in our questionnaire based on the results we obtained from our sample with a determined cutoff value of (≥0.7) (See Table 2). A comparison between male and female students was done using an independent sample T-test to identify if there were any statistically significant differences in response between each gender respectively with a determined cutoff value of (p≥0.05) (See Table8). Finally, a confirmatory factor analysis was done to test the reliability of our questionnaire on our targeted population based on the results we obtained from our sample with an acceptable cutoff values of root mean squared appropriation (RMSEA<0.08), standardized root mean squared residual (SRMR<0.08), Comparative fit index (CFI≥0.95), and Tucker-Lewis index (TLI≥0.95) (See Table 5);preceded by a KMO (Kaiser-Meyer-Olkin Measure) sampling adequacy test with an acceptable cutoff value of (>0.6) (See Table3) and a Spearman’s rho correlation test for the involved items (See Table 9) to eventually calculate the average variance extracted with an acceptable cutoff value of (p≥0.05) and the composite reliability with an acceptable cutoff value between (0.6-0.9) for each category respectively.(See Table 4). [12]

**Table 2:**
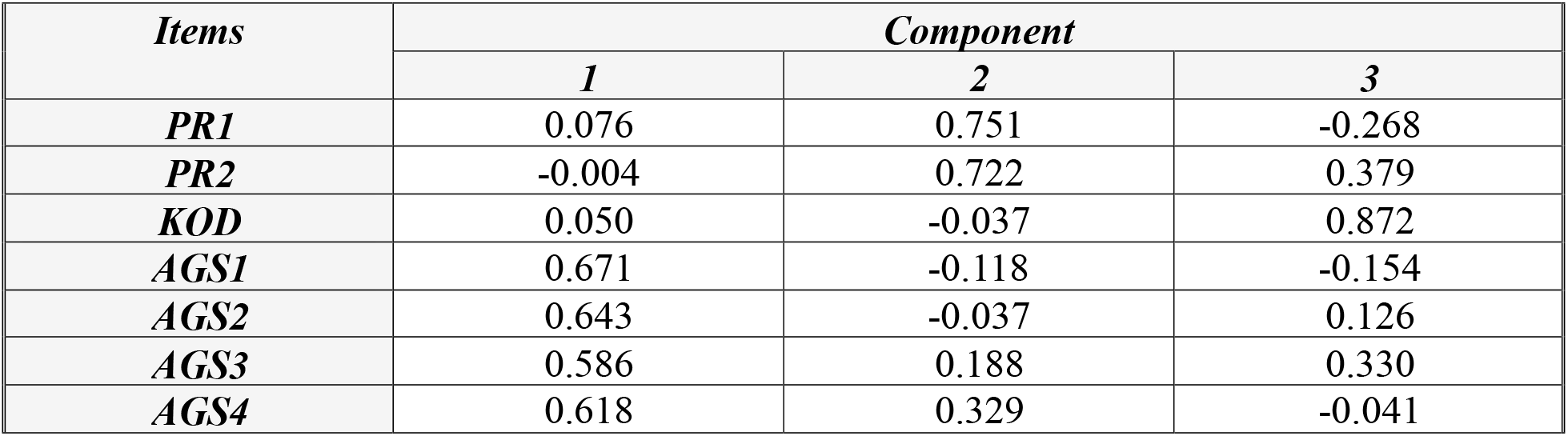
Exploratory factors analysis.

| *Items* | *Component* |  |  |
| --- | --- | --- | --- |
|  | *1* | *2* | *3* |
| *PR1* | 0.076 | 0.751 | -0.268 |
| *PR2* | -0.004 | 0.722 | 0.379 |
| *KOD* | 0.050 | -0.037 | 0.872 |
| *AGS1* | 0.671 | -0.118 | -0.154 |
| *AGS2* | 0.643 | -0.037 | 0.126 |
| *AGS3* | 0.586 | 0.188 | 0.330 |
| *AGS4* | 0.618 | 0.329 | -0.041 |

**Table 3:** Kaiser-Meyer-Olkin Measure of Sampling Adequacy.

|  |  |  |
| --- | --- | --- |
| *Kaiser-Meyer-Olkin Measure of Sampling Adequacy.* |  | *0.644* |
| *Bartlett's Test of Sphericity* | *Approx. Chi-Square* | *55.329* |
|  | *df* | *21* |
|  | *Sig.* | *0.000* |

**Table 4:** Confirmatory factor analysis 1.

| *Category* | *Average Variance Extracted* | *Composite Reliability* |
| --- | --- | --- |
| *Pattern Recognition* | *0.542* | *0.703* |
| *Knowledge of discipline* | *Non-applicable* |  |
| *Application of general strategies in problem solving* | *0.396* | *0.724* |

**Table 5:** Confirmatory factor analysis 2.

|  |  |
| --- | --- |
| **Comparative Fit Index (CFI)** | **1.000** |
| **Tucker-Lewis Index (TLI)** | **1.152** |
| **RMSEA** | **0.000** |
| **SRMR** | **0.012** |

**Table 6:** Descriptive statistics of the Three Categories and seven items of the problem solving skills model.

| **Category** | **Items** | **Mean** | **Standard deviation** |
| --- | --- | --- | --- |
| **Knowledge of discipline** | (1) I easily remember concepts that I learned long ago. | 2.973 | 0.999 |
|  | **Category total** | 2.973 | 0.999 |
| **Pattern recognition** | (2) I acknowledge my strengths and weaknesses on a particular topic. | 2.106 | 0.694 |
|  | (3) I recognize how much information I have about a concept. | 2.291 | 0.762 |
|  | **Category total** | 2.198 | 0.559 |
| **Application of general strategies for problem solving** | (4) I seek alternative solutions before making a decision. | 2.185 | 0.803 |
|  | (5) I decide the solution of a problem from evaluating several options. | 2.158 | 0.749 |
|  | (6) I identify alternative solutions to problems. | 2.158 | 0.712 |
|  | (7) I am able to focus on a problem. | 2.132 | 0.813 |
|  | **Category total** | 2.158 | 0.492 |

**Table 7:** Frequencies of the seven items.

| ***Likert scale*** | ***KOD*** | ***PR1*** | ***PR2*** | ***AGS1*** | ***AGS2*** | ***AGS3*** | ***AGS4*** |
| --- | --- | --- | --- | --- | --- | --- | --- |
| ***Strongly agree*** | 12 | 28 | 23 | 29 | 23 | 23 | 34 |
| ***Agree*** | 33 | 80 | 66 | 72 | 89 | 86 | 69 |
| ***Neutral*** | 61 | 42 | 57 | 44 | 32 | 37 | 43 |
| ***Disagree*** | 37 | 1 | 5 | 5 | 6 | 5 | 4 |
| ***Strongly disagree*** | 8 | 0 | 0 | 1 | 1 | 0 | 1 |

**Table 8:** **Descriptive statistics of the seven items of the problem solving skills model according to gender and independent sample T-test P value results.**

| *Items* | *Males* |  |  | *Females* |  |  | *P value* |
| --- | --- | --- | --- | --- | --- | --- | --- |
|  | Number | Mean | Standard of Deviation | Number | Mean | Standard of Deviation |  |
| **KOD** | 71 | 2.788 | 0.908 | 80 | 3.137 | 1.052 | **0.030** |
| **PR1** | 71 | 2.042 | 0.705 | 80 | 2.162 | 0.683 | 0.291 |
| **PR1** | 71 | 2.1972 | 0.785 | 80 | 2.375 | 0.735 | 0.155 |
| **AGS1** | 71 | 2.154 | 0.821 | 80 | 2.212 | 0.791 | 0.663 |
| **AGS2** | 71 | 2.154 | 0.821 | 80 | 2.162 | 0.683 | 0.951 |
| **AGS3** | 71 | 2.084 | 0.691 | 80 | 2.225 | 0.728 | 0.228 |
| **AGS4** | 71 | 2.098 | 0.813 | 80 | 2.162 | 0.818 | 0.632 |

**Table 9:** Correlations between the seven items.

|  |  |  | PR1 | PR2 | KOD | AGS1 | AGS2 | AGS3 | AGS4 |
| --- | --- | --- | --- | --- | --- | --- | --- | --- | --- |
| **SPEARMAN'S RHO** | **PR1** | Correlation coefficient | 1.000 | 0.142 | -0.005 | 0.070 | 0.115 | 0.015 | 0.113 |
|  |  | P value |  | 0.082 | 0.952 | 0.394 | 0.161 | 0.854 | 0.166 |
|  |  | Number | 151 | 151 | 151 | 151 | 151 | 151 | 151 |
|  | **PR2** | Correlation coefficient | 0.142 | 1.000 | 0.157 | 0.009 | 0.060 | 0.176 | 0.174 |
|  |  | P value | 0.082 |  | 0.054 | 0.912 | 0.464 | 0.030 | 0.032 |
|  |  | Number | 151 | 151 | 151 | 151 | 151 | 151 | 151 |
|  | **KOD** | Correlation coefficient | -0.005 | 0.157 | 1.000 | -0.008 | 0.081 | 0.145 | 0.074 |
|  |  | P value | 0.952 | 0.054 |  | 0.918 | 0.324 | 0.075 | 0.369 |
|  |  | Number | 151 | 151 | 151 | 151 | 151 | 151 | 151 |
|  | **AGS1** | Correlation coefficient | 0.070 | 0.009 | -0.008 | 1.000 | 0.256 | 0.156 | 0.195 |
|  |  | P value | 0.394 | 0.912 | 0.918 |  | 0.002 | 0.057 | 0.017 |
|  |  | Number | 151 | 151 | 151 | 151 | 151 | 151 | 151 |
|  | **AGS2** | Correlation coefficient | 0.115 | 0.060 | 0.081 | 0.256 | 1.000 | 0.247 | 0.204 |
|  |  | P value | 0.161 | 0.464 | 0.324 | 0.002 |  | 0.002 | 0.012 |
|  |  | Number | 151 | 151 | 151 | 151 | 151 | 151 | 151 |
|  | **AGS 3** | Correlation coefficient | 0.015 | 0.176 | 0.145 | 0.156 | 0.247 | 1.000 | 0.317 |
|  |  | P value | 0.854 | 0.030 | 0.075 | 0.057 | 0.002 |  | 0.000 |
|  |  | Number | 151 | 151 | 151 | 151 | 151 | 151 | 151 |
|  | **AGS 4** | Correlation coefficient | 0.113 | 0.174 | 0.074 | 0.195 | 0.204 | 0.317 | 1.000 |
|  |  | P value | 0.166 | 0.032 | 0.369 | 0.017 | 0.012 | 0.000 |  |
|  |  | Number | 151 | 151 | 151 | 151 | 151 | 151 | 151 |

## Results

The number of sixth Grade medical students in University of Baghdad/ College of medicine that participated in the study was 151 of whom 70 (46.35 %) were males and 81 (53.65 %) were females.

Regarding reliability, our questionnaire passed the kmo adequacy test scoring (0.644>0.6), and our confirmatory factor analysis results came all in the acceptable ranges [See Table 3]. Our RMESA (0.00013<0.08), SRMR(0.012<0.08), CFI(1≥0.95), TLI(1.52≥0.95) meaning our questionnaire is reliable on the application of general strategies for problem solving [See Table 4].The average variance extracted and composite reliability both were within the acceptablerange for each of the two categories with more than one item which in our case are knowledge of discipline and pattern recognition [See Table 5]

The mean score of the “knowledge of discipline” was 2.973± 0.999, as for the “pattern recognition” component the mean was 2.198 ±0.559, and the mean for the “application of general strategies” component was 2.158± 0.492.See (Table 6)

As for the individual items for (KOD) “I easily remember concepts that I learned long ago” 12 respondents answered strongly agree, 33 answered agree, 61 answered neutral, 37 answered disagree and 8 respondents answered strongly disagree, the mean for this item was. For the item (PR1) “I acknowledge my strengths and weaknesses on a particular topic” 28 respondents answered strongly agree, 80 answered agree, 42 answered neutral, 1 answered disagree and no one answered strongly disagree, the mean for this item was. For the item (PR2) “I recognize how much information I have about a concept” 23 respondents answered strongly agree, 66 answered agree, 57 answered neutral, 5 answered disagree and no one answered strongly disagree. For the item (AGS1) “I seek alternative solutions before making a decision” 29 respondents answered strongly agree, 72 answered agree, 44 answered neutral, 5 answered disagree, 1 respondents answered strongly disagree. For the item (AGS2)”I decide the solution of a problem from evaluating several options” 23 respondents answered strongly agree, 89 answered agree, 32 answered neutral, 6 answered disagree, 1 respondent answered strongly disagree. For the item (AGS3) “I identify alternative solutions to problems” 23 respondents answered strongly agree, 86 answered agree, 37 answered neutral, 5 answered disagree and no one answered strongly disagree. For the item (AGS4) “I am able to focus on a problem” 34 respondents answered strongly agree, 69 answered agree, 43 answered neutral, 4 answered disagree, 1 respondents answered strongly disagree. See (Table 6) and (Table 7)

For the genders males respondents were 71 and females were 80 and the results for the seven items were as follows: (PR1) “I acknowledge my strengths and weaknesses on a particular topic” had a mean of 2.042±0.705 for males and a mean of 2.162±0.683 for females, (PR2) “I recognize how much information I have about a concept” had a mean of 2.197 and a standard deviation of 0.785for males and a mean of 2.375±0.735, (KOD) “I easily remember concepts that I learned long ago” had a mean of 2.788±0.908 for males and a mean of 3.137±1.052,, AGS1 “I seek alternative solutions before making a decision” had a mean of 2.154±0.821 for males and a mean of 2.212±0.790 for females, (AGS2) “ I decide the solution of a problem from evaluating several options” had a mean of 2.154±0.821 for males and a mean of 2.162±0.683 for females, (AGS3) “I identify alternative solutions to problems” had a mean of 2.0845±0.691 for males and a mean of 2.225±0.728 for females, (AGS4) “I am able to focus on a problem” had a mean of 2.098±0.813 for males and a mean of 2.162±0.818 for females. See (Table 8)

Independent-Samples T test was used to observe any significant difference between male and female respondents for the seven items (questions) the values were as follows; (PR1) “I acknowledge my strengths and weaknesses on a particular topic” had P value of 0.291< 0.05 so no significant difference between male and female,(PR2) “I recognize how much information I have about a concept” had P value of 0.155<0.05 so no significant difference between male and female, (KOD)”I easily remember concepts that I learned long ago” had P value of 0.03>0.05 so there is a significant difference between male and female,(AGS1) “I seek alternative solutions before making a decision” had P value of 0.663<0.05 so no significant difference between male and female, (AGS2) “ I decide the solution of a problem from evaluating several options” had P value of 0.951<0.05 so no significant difference between male and female, (AGS3) “I identify alternative solutions to problems” P value of 0.226<0.05 so no significant difference between male and female, .(AGS4) “I am able to focus on a problem” had P value of 0.632<0.05 so no significant difference between male and female. Six of the seven items (PR1, PR2, AGS1, AGS2, AGS3, AGS4) had a P value of less than 0.05 so no significant difference between male and female and thus accepting the null hypothesis, these items of problem solving model are independent of gender. Except for one item (KOD) Had a p value of 0.030> 0.05 so there is a significance and thus rejecting null hypothesis and accepting alternative hypothesis this item is dependent on gender. See (Table 8).

Correlations between the seven items were calculated using Spearman’s rho Correlation coefficient and the results were: (PR1) had a Correlation coefficient of 0.142 with (PR2) with a P value of 0.082<0.05 so no significant correlation, and a Correlation coefficient of -0.005 with (KOD) with a P value 0.952<0.05 so no significant correlation, and had a Correlation coefficient of 0.070 with (AGS1) with a P value of 0.394<0.05 so no significant correlation, and had a Correlation coefficient of 0.115 with (AGS2) with a P value of 0.161<0.05 so no significant correlation, and had a Correlation coefficient of 0.015 with (AGS3) with a P value of 0.854<0.05 so no significant correlation, and had a Correlation coefficient of 0.113 with (AGS4) with a P value of 0.166<0.05 so no significant correlation. As for (PR2) the item had a correlation coefficient of 0.157 with (KOD) with a p value of 0.054<0.05 so no significant correlation, and had a correlation coefficient of 0.009 with (AGS1) with a p value of 0.912<0.05 so no significant correlation, and had a correlation coefficient of 0.060 with (AGS2) with a p value of 0.464<0.05 so no significant correlation, and had a correlation coefficient of (0.176) with AGS3 with a p value of 0.030>0.05 so there is a significant correlation, and had a correlation coefficient of 0.174 with (AGS4) with a p value of 0.032>0.05 so there is a significant correlation. As for (KOD) the item had a correlation coefficient of -0.008 with (AGS1) with a p value of 0.918<0.05 so no significant correlation, and had a correlation coefficient of 0.081 with (AGS2) with a p value of 0.324<0.05 so no significant correlation, and had a correlation coefficient of 0.145 with (AGS3) with a p value of 0.075<0.05 so no significant correlation, and had a correlation coefficient of 0.074 with (AGS4) with a p value of 0.369<0.05 so no significant correlation. As for (AGS1) it had a correlation coefficient of 0.256 with (AGS2) with a p value of 0.002>0.05 so there is a significant correlation, and had a correlation coefficient of 0.156 with (AGS3) with a p value of 0.057<0.05 so no significant correlation, and had a correlation coefficient of 0.195 with (AGS4) with a p value of 0.017>0.05 so there is a significant correlation. As for (AGS2) it had a correlation coefficient of 0.247 with (AGS3) with a p value of 0.002>0.05 so there is a significant correlation. And lastly for (AGS3) it had a correlation coefficient of 0.317 with (AGS4) with a p value of 0.000>0.05 so there is a significant correlation. See (Table 9)

## Discussion

In this cross-sectional study which is conducted for sixth grade students of Baghdad medical college, 151 respondents’ answers were gathered from which 80 of them were females and 71were males.

The descriptive statistics includes the results for each item. (Table 6) shows the mean and standard deviation of each item. Items with means closer to strongly agree are 2, 4, 5, 6 and 7, which correspond to the application of general strategies for problem-solving category and pattern recognition

The mean for the “pattern recognition” category was 2.198 (meaning answers leaning more towards strongly agree) which means students rate themselves fairly when it comes to identifying the pattern of a problem, as presented by previous study one way to enhance the students pattern recognition is to use visual and verbal guidance in approaching a problem rather than merely viewing the sequence, in other terms by letting the students be face to face with the problem rather than simply reading it.[13]

The mean for the “Application of general strategies for problem solving” category was 2.158 (meaning answers leaning more towards strongly agree), it had the mean that was closest to the (strongly agree) which means students rate themselves the highest (compared to the two other categories) when it comes to their ability of applying what they learnt in dealing with a problem, to enhance this ability even more as proposed by one study is to follow a problem based learning curricula rather than the conventional one, the study found that students in the problem based learning curricula produced extensive elaborations using relevant biomedical information, which was relatively absent from the conventional curricula students.[14]

The independent sample T-test results for the “Knowledge of discipline” category showed that the only item for this category (KOD) “I easily remember concepts that I learned long ago” had a mean for females of 3.137 (leaning more towards strongly disagree than males) and a mean for males of 2.778 (more leaning towards strongly agree than females) with a p value of 0.030>0.05 which means there is a statistical significance between males and female when it comes to assessing their knowledge as we can see that males assess themselves in remembering concepts as better than their female counterparts, this however contraindicates other studies, as mentioned before knowledge relies greatly on memory and a study found that when it comes to memory females tend to outperform their male counterparts.[15]

Regarding the “Pattern Recognition” category the independent samples T-test values were follows: (PR1) “I acknowledge my strengths and weaknesses on a particular topic” had P value of 0.291< 0.05, (PR2) “I recognize how much information I have about a concept” had a P value of 0.155<0.05, there was no statistical significance in “Patter Recognition” category between males and females, as both of them seem to rate themselves somewhat comparably, this was concluded in another study as Namrata Upadhayay and Sanjeev Guragain showed in their work that males and females have comparable cognitive functions when it comes to pattern recognition.[16]

For the “Application of general strategies for problem solving” category the independent samples T-test results were: (AGS1) “I seek alternative solutions before making a decision” had P value of 0.663<0.05, (AGS2) “ I decide the solution of a problem from evaluating several options” had P value of 0.951<0.05, (AGS3) “I identify alternative solutions to problems” P value of 0.226<0.05, .(AGS4) “I am able to focus on a problem” had P value of 0.632<0.05, there was no statistical significance in this category between males and females, meaning when it comes to applying what have been learnt to deal with a problem males and females perform and act some what the same.

For correlations between the categories one of the items of the “Pattern Recognition” category had a positive correlation with a statistical significance with 2 items from “Application of general strategies for problem solving” category, meaning that students who rate themselves higher as being able to identify the pattern of a problem also rate themselves fairly when it comes to applying what they learnt in dealing with it.

In order to understand why there is correlation between “Pattern Recognition” category and “Application of general strategies for problem solving” category instead of ‘‘knowledge of discipline’’, we need to understand how human brain process the information it receives There is more than one way to solve a problem perhaps the hardest, slowest and most time-consuming is to attempt to reason out a solution from basic principles. Certainly the easiest & most efficient is to recognize that you have solved it before and recall the solution.

Studies of cognitive psychology and functional magnetic resonance imaging demonstrate two distinct types of processes when it comes to decision-making: a fast, pattern-recognizing, intuitive way of thinking (type 1) and a more logical, analytical, and conscious, but a high-effort way of thinking that is highly dependent on the well-defined structure of knowledge (type 2)[17]. See (Table 10).

**Table 10:** Principal Characteristics of Type1 and Type 2 Decision Making process. adapted from Croskerry P. A universal model of diagnostic reasoning. Acad Med 2009; 84:1022–1028 [18]

| **Cognitive style** | **Type 1**<br>**Heuristic . intuitive** | **Type 2**<br>**Systematic . analytical** |
| --- | --- | --- |
| Responsiveness | *Passive* | Active |
| Capacity | High | Limited |
| Cognitive awareness | Low | High |
| Automaticity | High | Low |
| Rate | Fast | Slow |
| Reliability | Low | High |
| Errors | Relatively common | Rare |
| Effort | Low | High |
| Emotional attachment | High | Low |

This has been termed ‘dual process theory’. Dual process theory describes how the human brain has two distinct ‘minds’ when it comes to decision-making. This is could be related to forms of cognition that are ancient and shared with other animals – where speed is often more important than accuracy – and ones that are recently evolved and distinctly human. [18]

Each ‘mind’ has access to multiple systems in the brain In support of this theory, some fascinating objective data are emerging to potentially support the dual process theory. There are now functional MRI data to support the existence of different cognitive patterns. Activation of right lateral prefrontal cortex is noted when a logical task is correctly performed and when subjects inhibit a cognitive bias (type 2 thinking), a finding supporting this area’s potential role in cognitive monitoring. In contrast, when logical reasoning was overcome by belief bias, activity was noted in the ventral medial prefrontal cortex, a region associated with affective processing (type 1). [19]

Finally, there is some evidence that type 2 processing requires more blood glucose, and that alterations of blood glucose can modulate the type of processing predominantly used. [20]

Psychologists estimate that we spend 95% of our daily lives engaged in type 1 thinking – the intuitive, fast, subconscious mode of decision-making. [21]

Imagine driving a car, for example; it would be impossible to function efficiently if every decision and movement were as deliberate, conscious, slow, and effortful as in our first driving lesson. With experience, complex procedures become automatic, fast, and effortless. The same applies to medical practice.

This could explain the reason why students’ results show a high correlation between pattern recognition and the application of general strategies instead of knowledge of discipline in solving problem-solving model. students see themselves filling the defect of their knowledge by relying on pattern recognition to solve problems by using type 1 thinking Regarding this study students’ knowledge of discipline shows the lowest mean toward the strongly agree and no correlation between knowledge of discipline and pattern recognition nor application of general strategies which means they appear to be less confident to use their knowledge Students can improve their use of knowledge to solve problem OR type 1 thinking students by learning how experts utilize their knowledge, compared to novices (medical students), experts have a body of knowledge, strategies, and experiences accumulated over many years. Therefore, an expert/physician may see a different world—one that is not available to the novice.

There is natural progression in knowledge structure as learner progress from novice to an intermediate to an expert. Each is dependent on the evolution of knowledge structure: starting with guessing based on reduced knowledge; hypothetical deductive (hypothesis to data— backward reasoning) based on dispersed and elaborated knowledge structures; scheme inductive (signs and symptoms to disease—forward reasoning) based on a hierarchical knowledge structure; and pattern recognition based on a scripted knowledge structure. Thus, the clinical reasoning strategy used is dependent on the knowledge structure available to the learner. Scheme inductive reasoning only occurs when students’ knowledge structure is highly organized. [22]

There is a drastic difference in efficiency and accuracy depending on whether hypothetical deductive, scheme inductive, or pattern recognition is used A study was undertaken to determine the relationship between reasoning strategy used and likelihood of diagnostic success. Twenty experts and 20 novices each solved 12 cases (3 each of the clinical presentations of dysphagia, elevated liver enzymes, nausea and vomiting, diarrhea).Each subject was asked to think aloud as they solved the case and two independent judges rated the reasoning strategy as hypothetical deductive, scheme inductive, or pattern recognition. Of course, experts significantly outperformed students. In addition, performance was also dependent on the difficulty of the cases within each clinical presentation. But more importantly, it was found that students or experts who used scheme inductive or pattern recognition were five times more likely to get the correct diagnosis compared to subjects who used hypothetical deductive reasoning. [23]

Since the same scheme is utilized for both the inquiry and the organization of the knowledge just acquired the problem solving process reinforces the retention in long term memory of the organization of the knowledge relevant to specific problem, consequently the knowledge is learned. Maximizing relevancy, and minimizing information overload. unlike the scheme driven the hypothetical deductive or’ search and scan’ involves continuous testing of number of hypothesis for an appropriate match if the correct diagnosis is not among the hypothesis generated an accurate match may not occur. [24]

Clinicians should use both type 1 and type 2 thinking, as both types are important in clinical decision-making. When encountering a familiar problem, clinicians can use pattern recognition to reach a working diagnosis or differential diagnosis quickly (type 1 thinking). When encountering a more complicated problem, they use a slower, analytical approach (type 2 thinking). Both types of thinking interplay – they are not mutually exclusive in the diagnostic process [25].

## Conclusion

Sixth year medical students in University of Baghdad/ college of medicine are better self-perceived in the pattern recognition and application of general strategies for problem-solving categories of the problem-solving model than the knowledge of discipline, males assess themselves in remembering concepts as better than their female counterparts, students who rate themselves higher as being able to identify the pattern of a problem also rate themselves fairly when it comes to applying what they learnt in dealing with it. Further studies with larger sample size are needed to evaluate the entirety of students from different stages and correlate with their progress throughout their education.

## Data Availability

All data produced in the present study are available upon reasonable request to the authors

